# Group 2 and 3 ABC-transporter-dependent capsular K-loci contribute significantly to variation in the estimated invasive potential of *Escherichia coli*

**DOI:** 10.1101/2024.11.22.24317484

**Authors:** Rebecca A. Gladstone, Maiju Pesonen, Anna K. Pöntinen, Tommi Mäklin, Neil MacAlasdair, Harry Thorpe, Yan Shao, Sudaraka Mallawaarachchi, Sergio Arredondo-Alonso, Benjamin J. Parcell, Jake David Turnbull, Gerry Tonkin-hill, Pål J. Johnsen, Ørjan Samuelsen, Nicholas R. Thomson, Trevor Lawley, Jukka Corander

## Abstract

The major opportunistic pathogen *Escherichia coli* is the largest cause of antimicrobial resistance (AMR) associated infections and deaths globally. Considerable antigenic diversity has been documented in Extra-intestinal pathogenic *E. coli* (ExPEC), however, phenotypic capsular (K) typing has fallen out of use. We assembled and curated an *in silico* capsular locus typing database for group 2 and group 3 K-loci from >20,000 genomes and applied it to carriage and disease cohorts to investigate capsule epidemiology and evolution. Several widely circulating K-loci have estimated odds ratios of >7 for being found in bloodstream infections compared with carriage. The relative invasive potential differed between lineages, and notably, even between subclades of the multidrug-resistant ST131 clone. Our investigation highlights several capsules and lineages that contribute disproportionately to invasive ExPEC disease, some of which are also associated with high levels of AMR. These results have generated new insights into capsule epidemiology and evolution and have significant translational potential, including improved ExPEC diagnostics, personalised therapy options, and the ability to build predictive regional risk maps by combining genomic surveys with demographic and patient frailty data.

## Introduction

Capsules are major virulence determinants in bacterial pathogens. They have many critical roles, including shielding bacteria from the immune system, influencing the ability to cause invasive infections and acting as a barrier to antimicrobials. Bacterial capsular polysaccharide antigens are often well-studied and used as a central component of disease surveillance and effective vaccine targets, including for *Streptococcus pneumoniae*, *Haemophilus influenzae* and *Neisseria meningitidis*^1^. Whilst *Escherichia coli* is the leading cause of bacterial bloodstream infections (BSIs) globally and imposes the highest burden of antimicrobial-resistant BSI-associated deaths^2^, the true prevalence of different capsular types in contemporary disease has not been determined in large, unbiased extra-intestinal (ExPEC) infection cohorts. Capsules are natural targets for translational research to develop new antimicrobials and vaccines, and there have been early *in vivo* successes for phage therapy in invasive ExPEC infections^3^.

Traditional phenotypic *E. coli* capsular typing using a set of antisera is labour-intensive and no longer in general use. Unlike the O and H antigens^4^, no genotypic method exists for typing the K capsular loci. *E. coli* K-loci have been classified into four groups based on whether they are *wzy*-dependent (groups 1 and 4) or *kps*-ABC-transporter dependent (groups 2 and 3) and by the genetic organisation of the capsule locus^5,6^. The ExPEC-associated group 2 (G2) and group 3 (G3) K-loci have conserved capsular polysaccharide (*kps)* genes in Regions 1 and 3, which flank the capsular-determining genes in Region 2, making them ideal candidates for *in silico* typing. A G2 and G3 *E. coli* typing scheme complements the considerable work on phenotypes and structures to stimulate further study of *E. coli* K-types and their epidemiology^7^, as has occurred with typing schemes in other species and across bacterial families.^8–12^

## Results

By investigating a collection of >20,000 high-quality *E. coli* genomes, we observed 90 unique G2 (n=68) and G3 (n=22) ABC-transporter-dependent capsular (K) loci, based on capsular locus gene presence-absence patterns. These were used to create an *in silico G2 and G3 E. coli* K-typing reference database, compatible with the species-agnostic bacterial capsular loci typing tool Kaptive^13^. Eighty percent of the known G2 and G3 phenotypic K-types are represented here by phenotype-genotype paired data, yet known phenotypes only represent 34% of the 90 K-loci observed to have unique capsular gene sets. Furthermore, for 11 K-loci without a known phenotype observed in Norwegian BSIs, we confirmed a K+ phenotype with a positive precipitate with Cetavlon and found them negative for known K antigens, representing putatively novel K-types. We K-locus typed assemblies from several systematic studies of infections, including: 1) large longitudinal genomic surveys of bloodstream infections (BSIs) from Norway^14^ and the UK^15–17, 2^) urinary tract infection (UTI) surveys from Norway and France^18, 3^) UTIs with specific resistance profiles from the USA^19^, and 4) two collections of invasive isolates from neonates collected in low and middle-income countries (LMICs).^20,21^ In addition, K-locus typing of assemblies from infant-mother gut metagenomic surveys in the UK^22,23^ and from traditional culture picks (France^24^) provided information on K-types in asymptomatic colonisation. Throughout this manuscript, we will report the inferred phenotypic K-type (e.g., K1) for a given K-locus (e.g., KL1) genotype when available, while the numbering starts at KL110 for unknown phenotypes.

### K-locus epidemiology

European BSIs^14–17^ are predominantly caused by strains with a G2 or G3 capsule (81.3%); among these, G2 dominated, and only a minority had G3 loci (3.5%). The vast majority of BSI isolates in phylogroups B2 (94.2%), D (84.1%), and F (94.6%) had G2 K-loci. The top five K-types in European BSIs were K1, K5, K52, K2, and K14 (Fig. 1). These common K-types accounted for over 50% of BSIs (UK^15–17^, Norway^14^) and UTIs (Norway^18^, France^18^). Whilst G2 and G3 K-loci were far less common in asymptomatic colonisation in Europe (55.4%)^22–24^, K1 and K5 were still amongst the most common colonising G2-G3 K-types. Notably, G2 and G3 loci were much less common (53.9%) in invasive neonatal infections in LMICs^20,21^ (Fig. 1), a stark contrast to the 95% of UK BSIs in children <1 year old that were G2 or G3.

**Figure 1.**
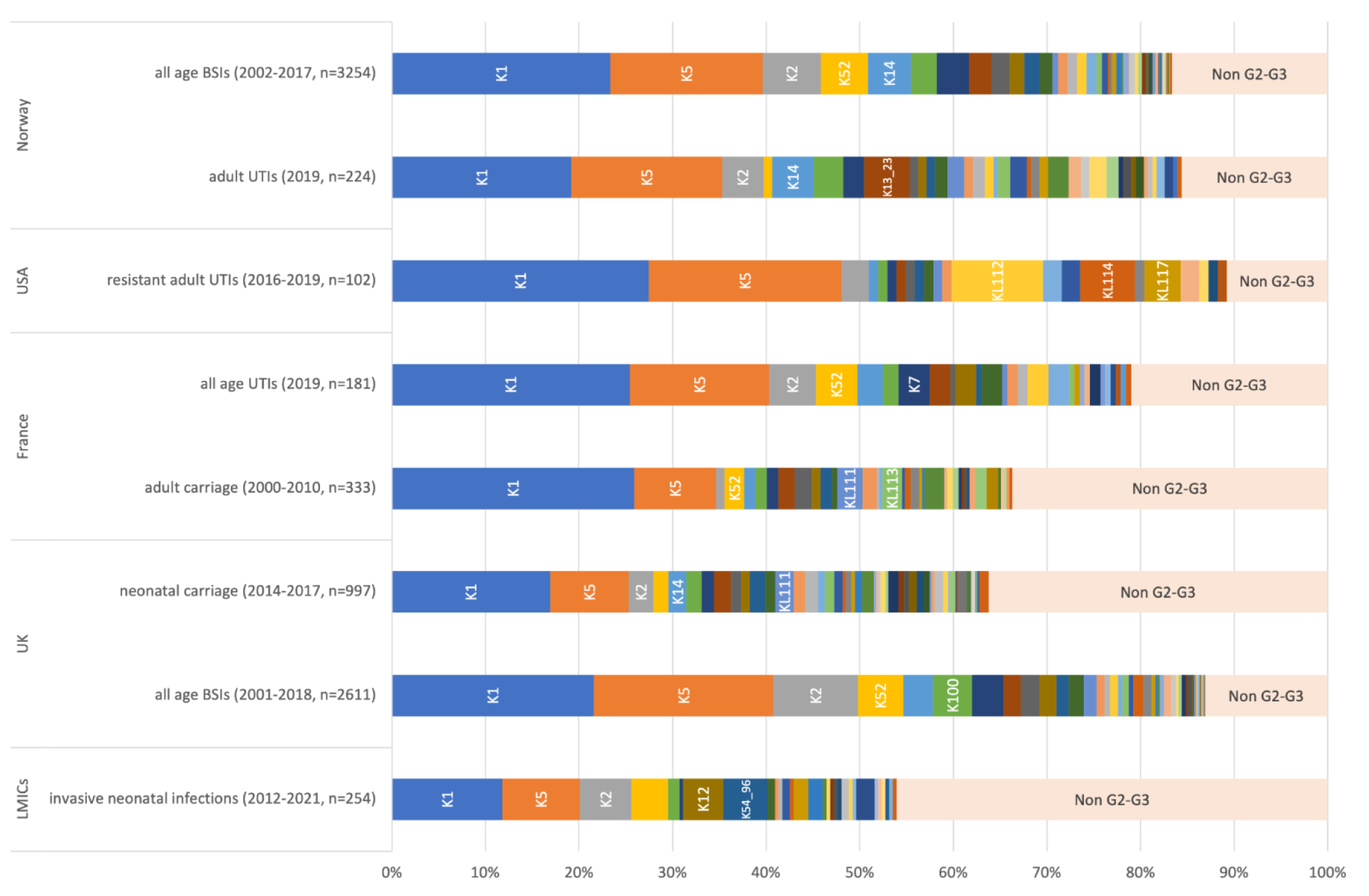
K-type prevalence across different collections. Published genomic collections were K-typed ^14–24^. The top 5 K-types in each collection are annotated. Untypeable isolates (*i.e.* genomes without G2-G3 *kps* genes) are classified as non-G2-G3.

### The estimated relative invasive potential of K-types

Using a mixed-effect model for UK isolates from carriage and BSIs, we estimated the contributions of each K-locus and lineage to the relative invasive potential (Figure 2). We determined that K52 had the highest invasive potential (OR=11.6, 95% CI: (5.3-25.4), p<0.0001) in comparison to the average untypeable isolate (genomes lacking kps genes). K14 and K100 had the second- and third-highest invasive potential, respectively (OR=8.9 [3.7-21.7], OR=7.2 [2.6-20.0]). The lineage-specific odds of K100 being found in BSIs were estimated to vary within CC131 between 5.42 (CC131 clade A) and 69.80 (CC131 clade C2). There was substantial variation in the lineage random effect estimates between CC131 subclades B and C despite sharing the same O-H type. The odds ratios for the widely studied K-types K1, K2, and K5 were ranked 12th, 5th, and 6th, respectively. These three types nevertheless had ORs significantly greater than 1, indicating greater invasive potential than untypeables. Conversely, K3 had the lowest invasive potential (OR=0.82 [0.2-2.7]).

**Figure 2.**
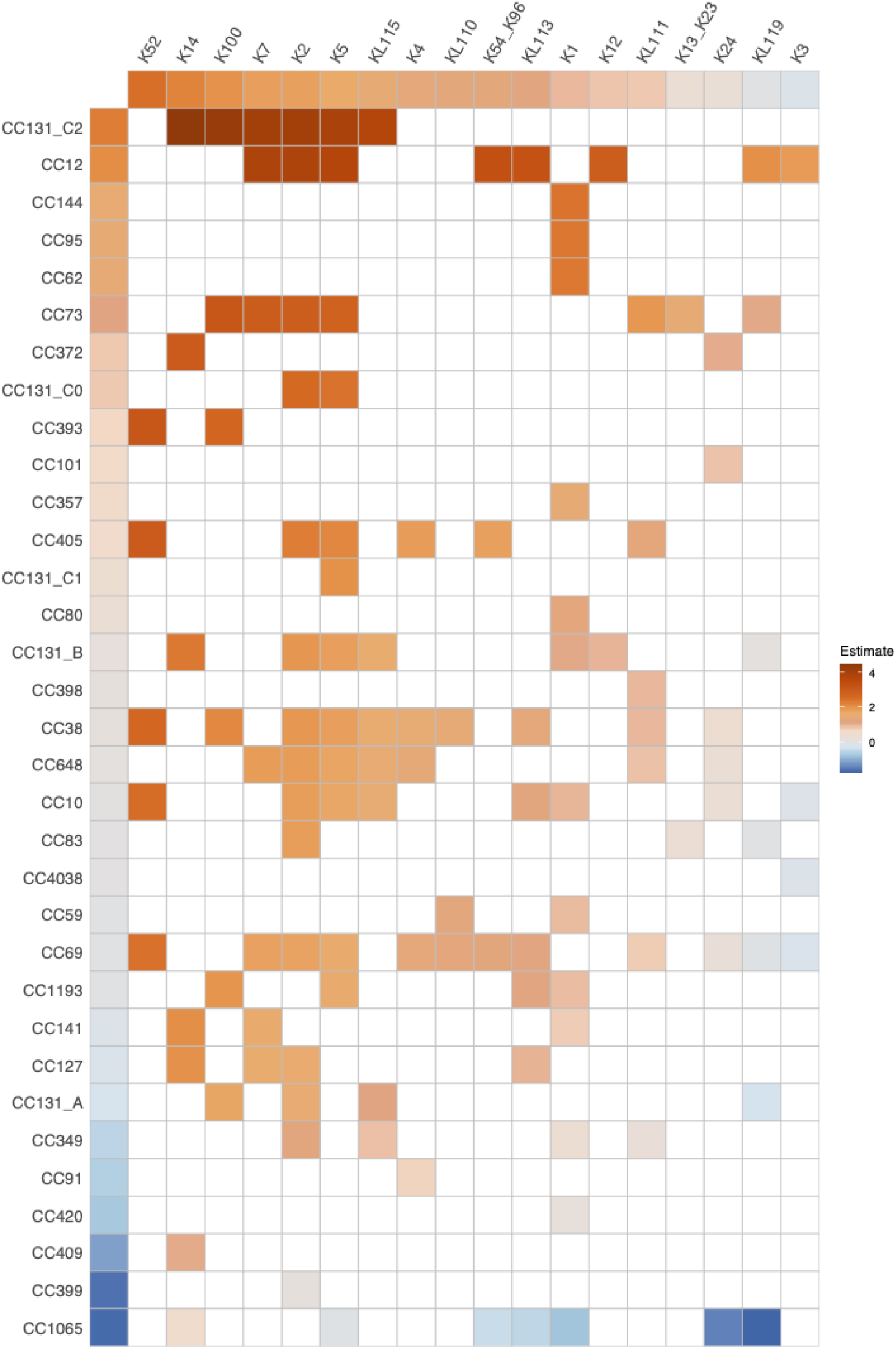
The estimated marginal and combined invasive potential of different K-loci (columns) and lineages (rows). The colours represent regression coefficients on a logit scale (log-odds) estimated from a generalised mixed model with clinical manifestation (infection/carriage) as a binary outcome, K-loci (KL) as a fixed effect and lineages as random effects. Lineages are denoted as Clonal Complexes (CC) derived from the representative sequence type for each popPUNK lineage. Red tones are associated with higher estimates of invasive potential, and blue tones with lower estimates. The reference category for K-loci was untypeable (*i.e.* genomes without G2-G3 *kps* genes). The rows and columns are sorted by the relative invasive potential estimates from highest to lowest. The CC131 lineage is split into its major clades (A, B, C1 and C2). The corresponding K-type, if known, is presented for a given K-locus.

We assessed the burden of E. coli BSI by age and sex and, in an exploratory analysis (Supplementary Figure 1), observed that K1 was overrepresented in the <1 and 40-49 age groups, but these differences were not significant after adjusting for multiple testing across all common age-K-type combinations. We observed that CC95-K1, but not K1 overall, was enriched in females (adjusted p=0.013), and K52, predominantly found in the uropathogenic CC69, was observed in females in every decade of life but only occurred in older men aged ≥50, making it significantly overrepresented in females (adjusted p<0.0001). The proportion of BSIs in the 1-59 age group was significantly higher among females (60%) than among males (40%; p<0.0001), but was similar overall (females 52%, males 48%). We quantified the odds of a BSI patient being >=60 years old in males compared with females for each common lineage-capsule combination. For most lineage-capsule combinations, there were no significant differences between males and females across the two age categories, except for CC131-C2 with the K5 capsule, where older males had higher odds. Limiting the model input data to only elderly adults (>=60, 74%) had a negligible effect on the rank order. We additionally determined that there was no correlation between the lineage random effects and the MDR or blaCTX-M proportion of that lineage (MDR R² = 0.029, p = 0.79, blaCTX-M (excluding C2) R² = 0.036, p = 0.74). Including the blaCTX-M proportion per lineage as a covariate in the model was not significant.

### Group 2 K-loci association with *E.coli* and the ExPEC pathotype

*E. coli* G2 Region 1 and 2 *kps* genes are not often found in other species. In a published collection of 661,000 assemblies^25^, only 11,737 were positive for the highly conserved and essential G2 *kpsF* gene (1kb) at 90% kmer identity. These were nearly exclusively *E. coli* (99.0%). Outside *E. coli*, *kpsF* was most commonly observed in *Salmonella enterica subspecies enterica* (n=69), *Klebsiella pneumoniae* (n=17) and *Staphylococcus aureus* (n=11). Using a published pangenome of ∼7,500 *E. coli*^26^, we assessed the pathotype association for G2. ExPECs accounted for 85.8% (1590/1853) of the *kpsF*-positive isolates with pathotype information, and non-ExPEC pathotypes accounted for 97.5% (3647/3741) of the *kpsF*-negative isolates. Shiga toxin-producing *E. coli* (STEC) accounted for the majority of non-ExPEC pathotypes positive for *kpsF* (90.5%, 238/263), and these STEC were found across multiple sequence types (ST), including ST442, ST10, ST25, ST504 and ST675.

### K-loci in major MDR lineages

The globally disseminated multidrug-resistant (MDR) lineage corresponding to clonal complex (CC)131 is known to have two dominant O-H types. However, we observed that at least 17 distinct K-loci were introduced into CC131, highlighting the rapid capsule diversification in this lineage (Fig. 3A). The ancestral K-type for the B/C clades is K5 (n=207/453) and K100 in clade A (n=80/104). Other K-loci with notable prevalence in CC131 BSIs include K2, K14, and KL112, all of which were estimated to have been acquired between 1980 and 2000. In total, K2 was acquired an estimated ten times independently across the lineage. Interestingly, KL112 in CC131 (n=27) is an example of a K-locus with an atypical architecture, where Region 1 is immediately followed by Region 3, and Region 2 is at the end of the locus. There were 5 subsequent fragmentation and complete G2 loss events after its acquisition.

**Figure 3.**
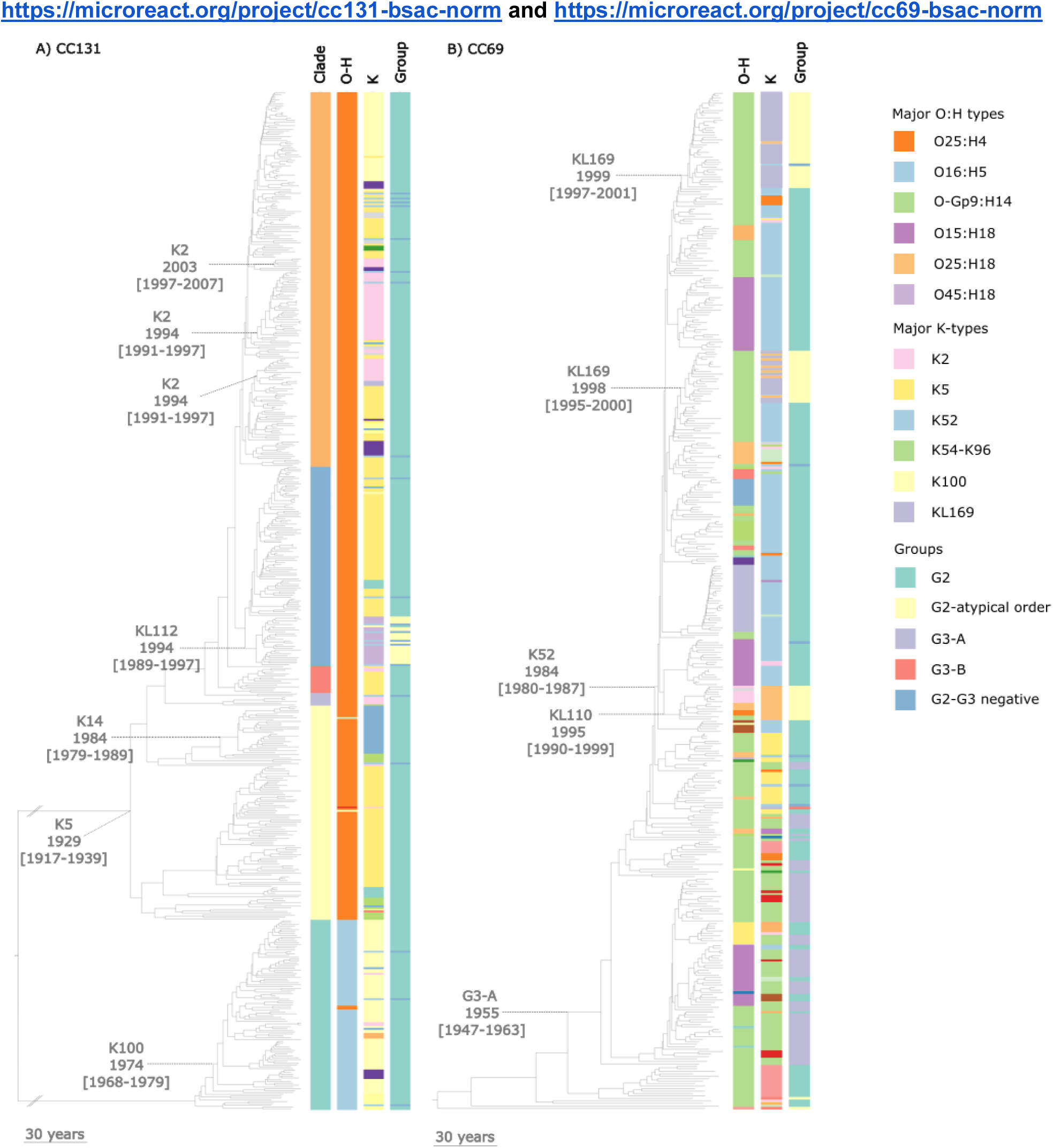
Diversity of K-loci across the clades of CC131 and CC69 from Norwegian and UK BSIs. Major capsule switch events are indicated with a dashed line from the relevant nodes to the time to the most recent common ancestor and confidence intervals. A) The four major CC131 clades are labelled in column one. The major O-H types are displayed in column 2 and the K-type in column 3. A split axis // is used on the branches leading from the root to clades A and B, as the root of CC131 was estimated to be nearly 600 years earlier than the A and B clades, this allows both trees to be viewed at the same time scale resolution B). The major CC69 O-H types are colour-coded in column 1, K-type in column 2 and the K-group in column 3. The adjacent key shows the major O-H and K-types. Where the K-phenotype is unknown, the K-locus (KL) is presented. Fully annotated interactive phylogenies are available at https://microreact.org/project/cc131-bsac-norm and https://microreact.org/project/cc69-bsac-norm

In another globally distributed MDR ExPEC lineage, CC69, we observed 11 O-types, 9 H-types and 23 K-types. O diversity in CC69 was higher than in CC131 (Simpson’s Diversity Index (SDI) CC69: 0.66, CC131: 0.39), but still less than CC69 K-locus diversity (SDI: 0.76). K-loci belonging to G2, atypical-G2 and G3 are all found in this lineage (Fig. 3B). The most common K-locus was K52 (39.5%, 163/413), which had the highest estimated invasive potential (Fig. 2). Switches from K52 to an unrelated atypical KL occurred at least three times since 1990.

### K-locus evolution

Whilst K diversity measured by SDI was higher than O or H diversity in over half of the lineages observed in BSIs, lineages varied greatly in their K-type diversity (Fig. 4A). The number of K-loci in a lineage was correlated with the r/m (recombination to mutation ratio) of that lineage, even when controlling for the lineage diversity by using the recombination-free median mutational pairwise distance (MMD, R2=0.95, p<0.0001, Fig. 4B). There was no correlation between r/m and MMD (R2=-0.35, p=0.3). The trend remained significant after excluding CC69 (R2=0.75, p=0.02), but not after excluding the two most recombinogenic lineages, CC69 and CC131 (R2=0.47, p=0.2). In the top four lineages in BSIs (CC73, CC95, CC69, and CC131), the *kps*-locus is a clear recombination hotspot, except for CC95, which expresses only K1.

**Figure 4.**
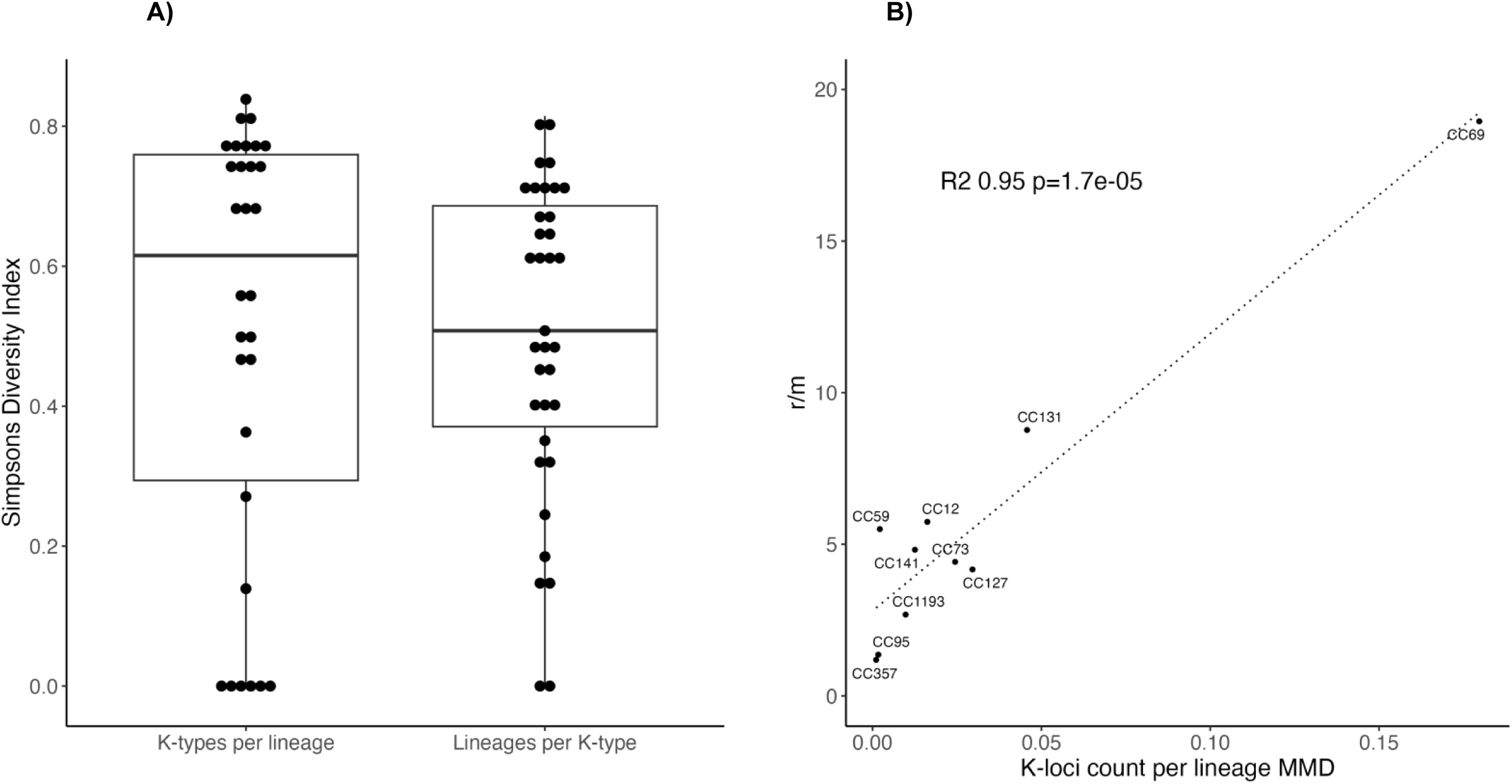
A) Simpson’s diversity index (1-D) for the richness and evenness of K-types within lineages, and of lineages within a K-type. B) Pearson correlation between the recombination to mutation ratio (r/m) and the number of K-loci per lineage, adjusted by the lineage diversity (median recombination-free mutational pairwise distance, MMD).

### K-loci structural variation

Known K-phenotypes accounted for only 34% of K-loci, with these phenotypes sporadically distributed across the K-loci phylogeny (Fig. 5). Many K-loci correspond to deep ancestral branches that likely represent distinct K-types. In total, there were 238 Region 2 capsular gene clusters (>70% amino acid AA sequence identity), of which 13.0% were annotated as “hypothetical protein”, demonstrating considerable unexplored capsular-determining gene variation. The eight *kps* genes in the G2 K-loci formed a single major gene cluster each. Only two of the G2 K-loci contained *kps* genes in a divergent cluster for at least one of the *kpsM, kpsT, kpsS* and/or *kpsC* genes, which are important in forming the biosynthesis-export complex^27^. Nine K-loci feature an atypical G2 locus structure that has not been previously reported, where Region 2 is outside of Regions 1 and 3 (Supplementary Figure 2). Although the atypical K-locus organisations are relatively rare in BSIs (<3.5%), they were spread across the species phylogeny and found in phylogroups A, B2, D and F. There were also sizable clusters of atypical loci in major *E. coli* BSI lineages: CC69 (KL110, n=84/449), CC131 (KL112, n=25/631), CC73 (KL112 n=20/981), CC59 (KL110 n=14/93). A Norwegian BSI isolate with the atypical K-locus KL144 was confirmed to be K+ but did not react with known K-antisera.

**Figure 5.**
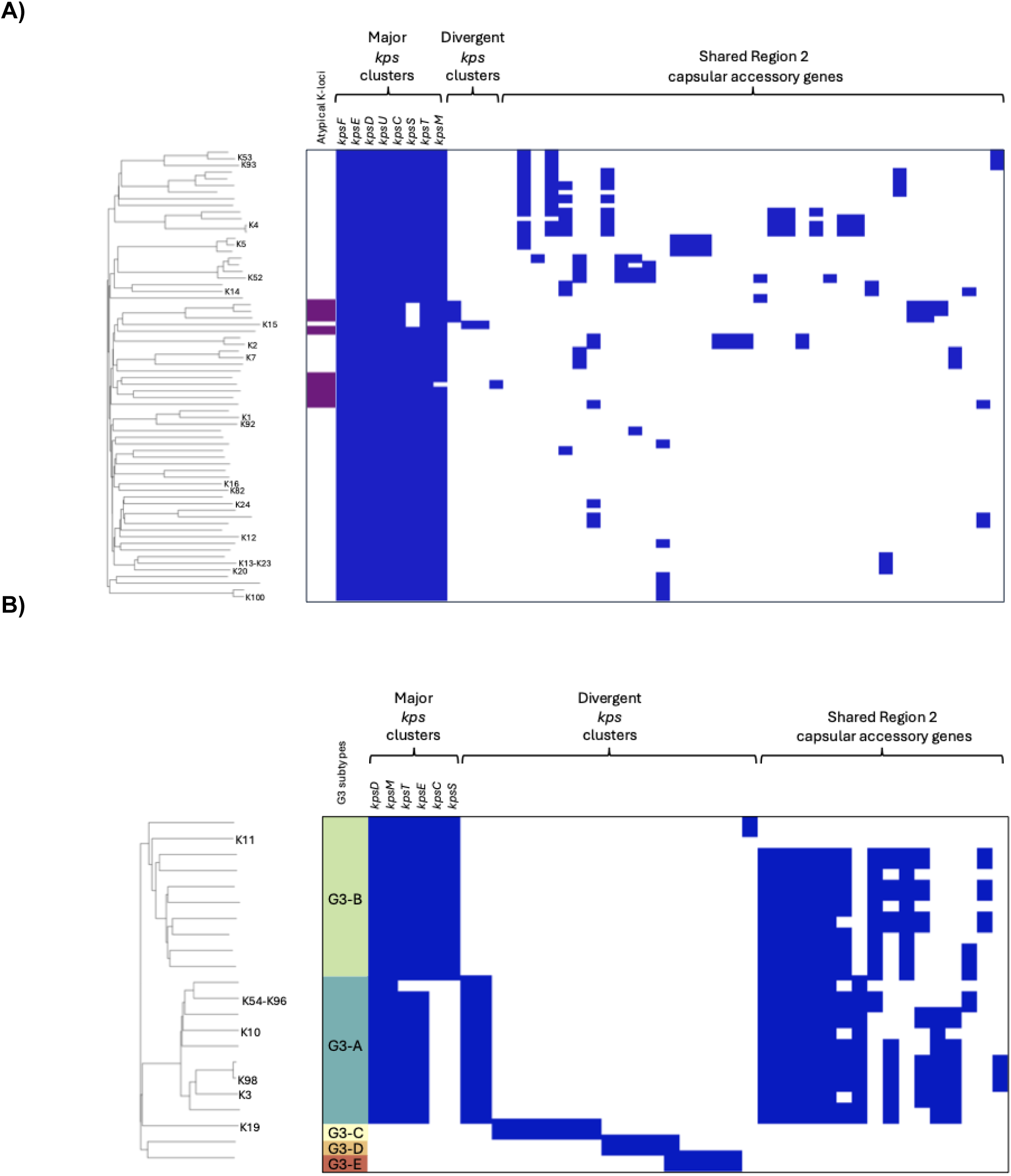
Neighbour-joining trees of G2 (5A) and G3 (5B) K-loci based on core and accessory sequence presence/absence, annotated with known K-phenotypes at the tips. The first metadata column denotes the K-loci with atypical gene organisation (5A) and subtypes of G3 (5B). The presence (blue) and absence (white) of gene clusters are shown. Three highlighted blocks show the dominant *kps* gene clusters in the order they are most often observed in the K-locus, divergent *kps* gene clusters, and the most common Region 2 gene clusters, ordered by the number of KLs in which they were observed. *only gene-clusters present in >2 K-loci are presented. These figures can be viewed interactively with full taxa and gene sets and annotation on phandango.net using the input files provided on https://github.com/rgladstone/EC-K-typing/tree/main/phandango

Unlike in G2, the diverse *kps* genes in G3 form two major gene clusters delineating the G3-A and G3-B subtypes (Fig. 5B). No G3 *kps* genes clustered with G2 *kps* genes. Within G3-A and G3-B the K-loci share Region 2 gene clusters for *wckE* and *rmlBDAC*. Meanwhile, K11, K19, KL151, KL160, and KL172 loci have divergent and unique Region 2 gene sets. K19 was observed in phylogroup A and shares no *kps* gene clusters with G2, KL172 has previously been classified as G3, and G3 has been suggested to be divergent G2 K-loci elsewhere^28,29^. Therefore, we denote these three loci as G3-C to G3-E.

### K-loci diversification and gene mobility

The proportion of unique K-loci sequences (at least 1bp difference) with one or more insertion sequences (IS) was 28.2% (1411/4996). While K1 was observed with an IS only once (n=1/692), K5 was observed with at least one IS in all cases (n=706). IS1 and IS3 were the most common of the ten IS families observed in the K-loci collection. Importantly, IS were observed overlapping with capsular coding sequences (CDS) in 17 K-loci and in nine of these, the IS carried a putative capsule gene in Region 2 as cargo.

The K-locus KL5 (K-type K5) is one of three closely related K-loci based on gene presence-absence patterns. The KL153 locus differs from KL5 by the loss of *kfiD*; it was rare and only observed in carriage (n=3). In KL127, an IS1 316 element contained a putative glycosyl-transferase family two protein (group 145) and a truncated *kfiC* within its terminal inverted repeats (TIRs, Supplementary Figure 3A) that was observed in BSIs (n=4) and carriage (n=5). An IS variant of the KL5 locus (KL5-1) seen in a single ST131 isolate had an additional tranposase and contained a fragment of the extended-spectrum β-lactamase gene *bla*_CTX-M_ near the IS3 remnant. The *bla*_CTX-M_ and transposase gene were observed together in multiple plasmid types within CC131 (100% identity), suggesting that both genes may have moved into this K-locus from a plasmid via homologous recombination with the existing IS in these K-loci.

There were four different K-loci related to K4 (KL4) with differing gene presence-absence patterns (Supplementary Figure 3B). The *kfoA, kfoB, kfoC,* and *kfoD* genes are present in all five, whilst *kfoE* and *kfoF* (gene-cluster name *udg*) were absent in KL148 and KL156, respectively. KL4-1 is an IS variant of KL4 where *kfoB* is split into two fragments and *kfoD* is contained within an IS element. These IS elements were all observed between *kpsS* and *kfoC* and likely contributed to the different patterns of capsular-specific genes within Region 2 for this cluster of related K-loci. K4 (KL4) was the most common of these loci in BSIs (n=54). Whilst IS variants were rare, their existence provides insight into how capsular diversification and gene exchange may be driven by IS. No IS was observed in the atypical loci, suggesting that other mechanisms are also at play.

G3-B K-loci have been observed in plasmids, which could also act as a vehicle for capsular mobility^29^. As G3-B are rare in BSIs (n=11), we observed just two Norwegian hybrid assemblies with G3-B K11 in multireplicon plasmids, but not in the chromosome. One of the isolates had K11 mobilised on a plasmid with multiple resistance genes: *bla*_TEM-1_ *dfrA14, mph(A), sul2, aph(3’’)-Ib and aph(6)-Id*. Additionally, a single *E. coli* hybrid assembly from Oxfordshire (n=1/549) was typed as G3-B KL132 on the plasmid and not in the chromosome.

## Discussion

Extensive phenotypic and structural data have demonstrated considerable diversity in the *E. coli* capsular antigens. Systematic cataloguing of capsular genetic diversity in contemporary disease is essential to further our understanding of the epidemiology of K-types. Here, we have shown that the diversity of G2 and G3 K-loci found in BSIs far exceeds the number of the currently known G2 and G3 phenotypic capsular types. Furthermore, this diversity has major implications for the development of preventive strategies, with observed differences in K-type prevalence by isolation source and geographical location, as well as variation in the estimated invasive potential of K-types across different genetic backgrounds. We determined that G2 and G3 capsules are present in the majority of ExPEC infections across diverse settings. Specifically, K1 and K5 are the most common K-types found in UTIs and BSIs. Although these two types are also the most common G2/G3 K-types in LMIC neonatal disease, a significant fraction of these infections are caused by non-G2-G3. While existing LMIC data are limited, our results clearly suggest that capsular epidemiology differs significantly from that in high-resource settings for at least vulnerable neonates, necessitating prioritising future sampling efforts to generate region-specific, representative data similar to that for other pathogens^30–32^. *In silico* typing of the *wzy*-dependent group 1 and group 4 capsules is another critical knowledge gap for understanding ExPEC disease in resource-limited settings. Nonetheless, the G2-G3 K-loci repertoire observed in LMICs was well represented by our G2-G3 database.

The role of capsules in pathogenesis has long been appreciated across bacterial species^1,33^. Previous research has experimentally demonstrated the virulence associated with common *E. coli* capsular types (e.g. K1, K2, K5, K52, K92 and K100) using either human immunological assays or murine models^34–39^. An epidemiological approach that leverages measured frequencies in systematically collected colonisation and disease isolates to calculate an odds ratio indirectly captures the relationship between exposure and the subsequent manifestation of disease cases in a population^16,22,40^. These estimates aim to capture a fixed trait of the capsule to provide a ranking for prioritising inclusion in vaccines and modelling optimal polyvalent vaccine formulations^41–47^.

Efforts to quantify the invasive potential of different K-types have been hindered by the scarcity of *E. coli* colonisation data, which we addressed by leveraging microbiome data from a large-scale UK infant-mother cohort^22,23,48^. Using infant carriage to represent general colonisation is one limitation of this approach. However, as healthy full-term babies acquire *E. coli* from family members and their surroundings during their first year of life, as reflected in strain sharing between mother and baby^22,49^, this suggests that the isolates seen in the infant population likely represent the commensals circulating in the general population. Furthermore, we found no significant differences in the overall K-type composition between the UK infant-mother collection and French adults, despite differences in the sampling period (2014-2017 vs. 2010), sequencing approach (metagenomic vs. single-colony sequencing), and geographical location. The similarity of the two carriage populations suggests that there is limited age structure to human *E. coli* colonisation in this geographical region. Whilst a single capsule type and a clonal complex (K1 and CC10) had a significantly higher prevalence in the French collection, these differences did not affect our conclusion that high K1 carriage prevalence influences its high prevalence in disease.

Given the similarity between neonatal and adult carriage, we assumed that *E. coli* carriage is homogeneous across age groups. Whilst this carriage represents the general healthy population, the majority of BSIs occur in elderly adults who often have comorbidities. The estimates reported in this manuscript, therefore, reflect the capsular types that are enriched in BSI, as compared to the general healthy adult population. Colonisation in elderly adults may differ from that in the general population, and a matched cohort of colonised patients who did not progress to BSI would provide an ideal control to quantify the causal effects of capsules, accounting for potential age-based biases. We observed some age- and sex-related differences in BSI epidemiology, of note was an overrepresentation of men in the elderly adult category, suggesting that the invasive potential may differ across patient groups. However, we could not directly integrate age and sex as covariates into the model due to the strong correlation between age and dataset membership and the observed age-sex interaction in the risk analysis. We assessed this limitation in two ways: first, we found that the odds of a BSI patient being >=60 in males compared to females were not significant for most lineage-capsule combinations, except for CC131-C2 with K5. Secondly, as we could not explicitly include age as a covariate, we highlight that the elderly adult high-risk age group (>=60 years) is overrepresented in the data (74% of BSIs) and, therefore, that the estimates are most relevant for this important clinical population. Furthermore, we ran additional iterations of the model, excluding the other age groups, to remove the assumption that the invasive potential is homogeneous across age groups and limit it to the simpler assumption for which we have some evidence: that carriage is homogeneous in the healthy population. Limiting the data to only elderly adults had a negligible effect on the rank order but increased uncertainty in the confidence intervals of all effect estimates for the K-types. Whilst there was insufficient data from elderly adults alone to estimate the relative invasive potential of four capsular types, one notable difference was that K12 moved up four places in the rank order. It may represent a K-type that either requires a more vulnerable population or circulates more commonly in older demographics, which were not captured by the infant-mother carriage cohort.

Hard-to-treat infections may be overrepresented in BSI isolates due to a greater likelihood of progression to systemic infection, but this was not a significant covariate. To further assess the robustness of the model, we applied it to a pneumococcal collection previously used to assess invasive potential ^50^ and reproduced the general rank order of invasiveness reported by others, including capsules previously established as the most virulent for this bacterium^41,47,50^. As further colonisation data are generated alongside additional BSI metadata, there is potential for future meta-analyses across countries to increase power to estimate the relative invasive potential of K-types within different groups and reduce uncertainty around point estimates.

Our results suggest that the high frequency of the widely studied K1 and K5 types in disease is, at least in part, a product of greater opportunistic spillover into vulnerable hosts. Our estimates also indicate that multiple K-types are more strongly associated with infection when ranked by invasiveness. For example, K52 and K14, which are found in multiple lineages, including the global MDR lineages CC69 and CC131, had the highest estimates of invasiveness and subsequently ranked 3rd and 4th in BSI prevalence despite rarely being seen in carriage samples in the UK and France (<2.5%). Whilst the estimated invasive potential of K1 was not top-ranked, the ST95 lineage ranked 4th in the lineage random effects estimates, suggesting the lineage is playing a key role in progression to bloodstream infections, which is concordant with the many reported virulence factors in this lineage^51–54^. In general the lineage estimates are of similar magnitude to K-types at least for the top ranked lineages, again suggesting that other virulence factors within a lineage encoded outside the K-locus are equally important for determining invasiveness.

The development of ExPEC vaccines is complicated because many *E. coli* are commensal, and some are considered beneficial to human health^55^. Vaccines should avoid targeting a major constituent of the gut microbiota and broad *E. coli* sterilising immunity. This requires a greater understanding of the differences between predominantly commensal genotypes and successful ExPECs, as well as the pathogenic potential of the latter. K-types with high estimated invasive potential are obvious vaccine targets due to both their capacity to cause disease and the likelihood that removing them from the gut flora will less often affect key *E. coli* commensals or lead to large-scale replacement in carriage and disease as other colonisers fill the vacated niche. While ExPEC vaccines have been under development since the 1980s, only a few have been licensed. Whole-cell, O, H, K, and O-conjugates have all undergone clinical trials^56^.

Although evidence of K1 and K5 autoreactivity is limited, their mimicry of host glycobiology may explain why these anti-K antibodies are not strongly induced in vaccination or disease^56^. Nonetheless, increased genomic data and expanded O-typing have contributed to the increased development of subunit-based vaccines rather than whole-cell vaccines in recent years^4,57^. Currently, there are over twice as many recognised O-serogroups as K-types^56^. Yet, we observed a greater diversity of K-loci from G2 and G3 than O-loci in most dominant ExPEC lineages.

Even greater diversity is likely to be discovered with further investigation into the genetic determinants and delineation of serogroups. As little as a single base pair difference has been seen in capsule-determining glycosyl-transferases of other species^58^. Indeed, we observe multiple serotypes with the same capsular genes with high sequence identity. These include the K13 and K23 reference phenotypes, both typed as KL13. They have previously been reported to belong to a serogroup, along with K20, which has one gene replaced relative to K13 and K23^59^. The G3 phenotypes K54 and K96 also share a set of capsular genes and are typed as KL96^60^. Clusters of K-loci that differ by a single Region 2 gene or by gene truncations could also represent additional cross-reactive serogroups. Alternatives to traditional K-phenotyping are in development that could discriminate between subtly different K-phenotypes and allow novel K-types to be assessed more readily.^61^ The putative genetic determinants need in-depth phenotypic validation to allow full genomic discrimination, which could be included as phenotype logic to Kaptive3 ^62^.

The evolution of the *E. coli* G2 and G3 K-locus is of considerable interest, given the fundamental biological role of the capsule and the complexity of its biosynthesis. We observed that a specific subgroup of G2 K-loci was undergoing atypical diversification, one of which had an invasiveness OR significantly above 1, indicating that atypical K-loci can be clinically important. We demonstrated that the overall tendency of homologous recombination across the core chromosome closely reflects the rate at which capsule switches occur via horizontal gene transfer, thereby influencing the K locus. Whilst earlier work detected the acquisition of the K1 capsule by genetic lineages across several phylogroups over centuries^36^, here we demonstrate that a similar process has influenced the spread of most of the successful and invasive capsule types. The ability to diversify the capsule locus for two major MDR lineages (CC69 and CC131) may have facilitated their rapid expansion in BSIs in the 21st century.^14–16^ We further discovered that IS elements are common within the K loci and are likely contributing to the diversification of the polysaccharide composition of capsules by importing genes into Region 2, warranting deeper functional investigation of these evolutionary processes. This appears to be a unique feature of *E. coli* G2 and G3 capsules; whilst IS-elements have been reported in other species like *Klebsiella pneumoniae*^8,9^, they could be completely removed from the typing scheme as they were less common and purely intergenic.

In summary, our results provide impetus for reinstating phenotypic capsular typing for this species and renewed interest for experimental studies of virulence that could further disentangle the contributions of capsules versus lineages towards invasiveness, to allow the development of strategies to effectively control ExPEC disease.

## Methods

### Data

We used a published pangenome analysis of ∼7,500 high-quality *E. coli* genomes to determine that the capsular gene *kpsF* was consistently annotated and predictive of G2 capsule presence^26^. We subsequently downloaded all *kpsF*-positive (0.9 kmer-ID threshold) *E. coli* from a published, searchable collection of 661,000 bacterial assemblies (n=11,623)^25^. For G3, we additionally screened that database for all *kpsM*-positive assemblies (0.9 kmer-ID threshold, n=853) using the G3-A and G3-B *kpsM* alleles from K96 and K11, respectively, as *kpsM* is more divergent in G3. We supplemented these genomes with data from large published *E. coli* BSI longitudinal genomic collections from Norway (2002-2017, n=3,254, 60% hybrid assemblies)^14,63^ and the UK (2001-2018 n=2,219)^16^, and a collection of infant and mother *E. coli* carriage assemblies from the UK babybiome study (n=997, available from https://zenodo.org/records/14000489)^22,23^, and two one-health *E. coli* studies from the UK (n=405) and USA (n=2948)^64,65^. Genomes with a known K phenotype were sourced for 24 different K-types from GenBank, the NCTC project^66^ and Enterobase^67^. The NCTC reference strains (accession PRJEB6403) are maintained as preserved strains that can be obtained from the National Collection of Type Cultures, UK. K-loci were initially extracted from 21,802 assemblies using an *in-silico* PCR (https://github.com/simonrharris/in_silico_pcr) for known K-loci boundaries, i.e. *kpsF*-*kpsM* or *kpsM/D-kpsS*. PCR-negative K-loci were then manually extracted from assemblies that had been annotated with one or more *kps* genes.

### K-loci

Extracted K-loci with unknown bases were excluded, and the K-loci were filtered down to 4,996 unique sequences with at least 1bp difference. These unique K-loci were annotated with Prokka^68^ and analysed with Panaroo^69^ to consistently annotate K-loci gene clusters with a conservative 70% family identity threshold. The gene cluster names were updated using the capsular-specific reference annotations to reflect the literature where possible. This resulted in an initial 225 gene absence patterns, excluding insertion elements (IS). IS elements were identified from the annotations using ISEscan^70^. These 225 sequences were pruned to remove K-loci with redundant patterns due to misannotation, paralogs, non-capsular genes, and incomplete K-locus remnants, leaving 90 curated K-loci in the final K-loci database. K-loci from complete genomes, long-read/hybrid assemblies and sequences containing the fewest IS were preferentially included as the database reference. The database was formatted for Kaptive version 3.0.0b5 ^13^, with IS annotations removed. Unpublished data from European disease (n=1,592) and carriage from underrepresented regions in Asia and Africa (n=10,085) were screened to determine that no dominant K-types had been missed. KL loci nomenclature reflects the known paired phenotypes, e.g. K1 is encoded by the KL1 locus. K-loci for which K-types have not yet been phenotypically identified were assigned KL numbers starting from KL110.

Assemblies (n=8,473) from published genomic collections ^15,16,18–20,22–24,71^ were K-typed using Kaptive, and our G2-G3 database version 3.0.0, which is available at https://github.com/rgladstone/EC-K-typing. Untypeables with three or fewer essential *kps* genes were designated absent for G2-G3 K-loci. The assignment of chromosome or plasmid was available for hybrid assemblies from Norway and Oxfordshire^17,63^. These were used to determine whether any K-loci were present in plasmids from these BSI collections.

The relatedness of K-loci was assessed for G2 and G3 separately using the pairwise mash hash distance of the K-locus sequences converted to a proportion 1-(n/1000), which gives greater resolution than gene presence-absence but still captures core and accessory variation^72^. These distances were used to create Neighbour-joining phylogenies in the R v4.4.1 package ape 5.8 and visualised in Phandango.net^73^ using the following input files https://github.com/rgladstone/EC-K-typing/tree/main/phandango.

### Phenotypic confirmation

One hundred and fifty Norwegian BSI isolates were selected for phenotypic K-typing; these were selected to represent imperfect matches to K-loci with known phenotypes (n=71), K-loci where a phenotypic reference genome was not available (n=40) and isolates which were *kps*-negative (n=39) to distinguish between G1-G4 and potentially acapsular isolates. K-phenotyping was carried out by the Staten Serum Institute using reference capsular antisera. These data were used to confirm phenotypes assigned in the K-typing database and to determine whether K-loci without a known K-phenotype expressed a putatively novel capsule, as evidenced by a positive precipitate reaction with Cetavlon but negative with known K antigens.

### *E. coli* carriage assemblies from metagenomic data

*E. coli* assemblies were extracted from metagenomic data from the UK babybiome data using a computational approach described in a previous study^74^ to provide an *E. coli* gut colonisation dataset^23,75^. The sequencing read data were first pseudo-aligned against a diverse reference database constructed from the 661,000 assemblies study^25^ (available from https://zenodo.org/records/7736981) with Themisto^76^. The alignments were then processed with mSWEEP and mGEMS^77,78^ to assign each sequencing read to a bacterial species. Next, the reads assigned to *E. coli* were pseudo-aligned with Themisto against an *E. coli* database (https://zenodo.org/records/12528310) and reprocessed with mSWEEP and mGEMS to obtain assignments of the reads to *E. coli* PopPUNK lineages^79^. The resulting assignments (‘bins’) were quality controlled with demix_check (https://github.com/harry-thorpe/demix_check) and bins with scores 1 or 2 were retained (n=1,402). Reads belonging to the 1,402 bins were assembled with Shovill (https://github.com/tseemann/shovill), and small contigs <5000bp were discarded before K-typing. The assemblies are available at https://zenodo.org/records/14000489. As this collection sampled individuals within a family (mother and infant) multiple times during the first year of life, the isolates were deduplicated (n=997/1,402), allowing only one lineage representative with the same K-type per family. Mothers accounted for 67/997 of the isolates, and infants between 6 months and 1 year old accounted for one third of the deduplicated set. When both a typeable and an untypeable isolate were observed within a family for a particular lineage, we preferentially selected the K-typed isolate, as it is likely to be of the same strain but with a higher-quality assembly; this accounts for the fact that lower-coverage assemblies limit K-type detection.

### Relative invasive potential

Lineages were defined based on core and accessory distances using PopPUNK^79^. Each lineage is named after the representative sequence type for that popPUNK cluster and described as a clonal Complex (CC), as each lineage contains multiple STs. To avoid a temporal bias whilst estimating the relative invasive potential, due to differences in collection years between BSI (2001-2017) and carriage (2014-2017), we excluded BSIs from 2001-2002 from the analysis to remove a known expansion of the CC69 and CC131 lineages, which later reached a stable equilibrium population frequency by 2003. Furthermore, we tested for differences in proportions for each lineage and K-type between the 2003-2013 and 2014-2017 periods using two-sided Fisher’s exact tests, and we adjusted for multiple testing. Only CC393 showed a significant difference between the periods, which could introduce bias. The K-loci, lineage information, and the source of each isolate (infection/carriage) were input into a generalised mixed model to estimate the relative invasive potential of K-types and lineages^15,16,22,23^. The model included the isolation source as a binary outcome variable, the K-loci indicator variable as a fixed effect (constant for each isolate) and the lineage indicator variable as a random effect. Untypeable (*kps*-negative genomes or rare loci) was set as the reference category for K-loci. Data were filtered to include only K-loci found in more than 20 isolates, with at least 5 isolates in each infection and carriage group. The model was fitted in R using the glmer function in the lme4 package (v1.1.35.5)^80^. The age and sex of BSI patients, the likelihood that a strain was CTX-M positive, and an increase in BSI prevalence of CC393 between 2002-2013 and 2014-2017 were possible confounders, the model was run five additional times: 1) BSAC restricted to adults only n=1550/2036, 2) BSAC elderly adults only n=1202/2036, 3) BSAC with CC393 subsampled in 2003-2013 down to the same proportion as 2014-2017 of 0.02 n=2008/2036, 4) adjusting for the CTX-M prevalence of each lineage in BSIs and 5) with sex as a covariate. We further assessed whether the infant-mother metagenomic carriage data had significantly different K-epidemiology to a healthy adult carriage whole genome collection ^24^ sampled in France in 2010, using a nonparametric test of independence in R package coin v1.4-3 for the K-loci and lineages included in the invasive potential model, and a comparison of proportions for each individual K-type and lineage included in our model between these two carriage collections with the “BH” method for adjusting for multiple testing. We computed ORs with 95% CIs and p-values for the top CC+K combinations with at least 50 isolates in the BSAC BSI collection to assess the age-sex interaction within BSIs. This allowed us to compare the odds of a BSI patient being >=60 years old in males versus females for each CC+K combination. The ORs, CIs and p-values were estimated with Fisher’s exact test. The model was also applied to a previously published pneumococcal dataset for assessing invasive potential^41,50^, leveraging the availability of paired carriage and disease collections from two countries in the model as a fixed effect. This multi-country approach increased the sample size to 1,415 carriage isolates and 1,081 disease isolates. https://github.com/rgladstone/EC-K-typing/tree/main/invasiveness/pneumo_example.

### Lineage analysis

The top 10 PopPUNK^79^ lineages were mapped against a corresponding ST reference, aligned, and recombination removed with Gubbins^81,82^. The recombination over mutation ratio (r/m) per lineage was estimated with Gubbins, and the median pairwise recombination-free mutational distance (MMD) for each lineage was calculated using pairSNP^83^. The Pearson correlation between r/m and the K-locus count per lineage, adjusted for lineage diversity using the MMD, was calculated in R v4.4.1 using base::corr.test for the top 10 lineages with >95% G2-G3 capsules. For dating analysis, recombination-free phylogenies for CC69 and CC131 using Norwegian and UK BSI data^14,16^ were created with Gubbins v3.4^82^ using SKA2^84^ and dated with the BactDating^85^ ARC model with a 10,000,000 MCMC chain in three replicates and one with randomised tip dates. The effective sampling size was >200, the Gelman statistic for convergence was ∼1, and the true date models were better than the random date models. The resultant phylogenies were visualised in microreact^86^. To characterise the diversity of K-loci within a lineage, we used Simpson’s diversity index (SDI 1-D), which measures the relationship between the number of “species” (here, either K-loci or lineages), termed richness, and the number of individuals within each “species”, termed evenness. O:H-typing was performed with SRST2^4^.

## Data availability

The genomic accession numbers and metadata that support the findings of this study are available in the Supplementary data and https://github.com/rgladstone/EC-K-typing.

## Correspondence and requests for materials

Corresponding authors: Rebecca A. Gladstone, Jukka Corander

## Acknowledgements

We thank our collaborators forming The Norwegian *E. coli* BSI Study Group: Nina Handal (Akershus University Hospital), Nils Olav Hermansen (Oslo University Hospital, Ullevål), Anita Kanestrøm (Østfold Hospital), Hege Elisabeth Larsen (Nordland Hospital), Paul Christoffer Lindemann (Haukeland University Hospital), Iren Høyland Löhr (Stavanger University Hospital), Åshild Marvik (Vestfold Hospital), Einar Nilsen (Molde Hospital and Ålesund Hospital), Marcela Pino (Oslo University Hospital, Rikshospitalet), Elisabeth Sirnes (Førde Hospital), Ståle Tofteland (Sørlandet Hospital), Kyriakos Zaragkoulias (Nord-Trøndelag Hospital Trust). We also thank Dr Kat Holt and Dr Kelly Wyres, who kindly advised us in maximising the database’s utility with the Kaptive tool. Finally, we thank Susanne Jespersen and Marie Vilborg Jacobsen for their efforts in K-typing 150 isolates supported by the Gates Foundation.

## Inclusion and ethics statement

This publication used published genomic collections and their associated metadata.

## Individual contributions

**RAG**: Conceptualization, Methodology, Validation, Formal analysis, Investigation, Data Curation, Writing - Original Draft, Visualization, Project administration, Funding acquisition **MP**: Methodology, Software, Formal analysis, Data Curation, Writing - Review & Editing, Visualization. **AKP**: Investigation, Writing - Review & Editing. **TM**: Data Curation, Formal analysis, Writing - Review & Editing. **NM**: Software, Writing - Review & Editing. **HT**: Writing - Review & Editing. **YS and SAA**: Data Curation, Writing - Review & Editing. **SM and GTH**: Methodology, Writing - Review & Editing. **BJ and JDT**: Resources, Data Curation, Writing - Review & Editing. **PJJ, NRT and TL**: Resources, Writing - Review & Editing. **ØS**: Resources, Validation, Writing - Review & Editing. **JC**: Conceptualization, Resources, Writing - Original Draft, Supervision, Project administration, Funding acquisition

## Supplementary

**Supplementary Figure 1.**
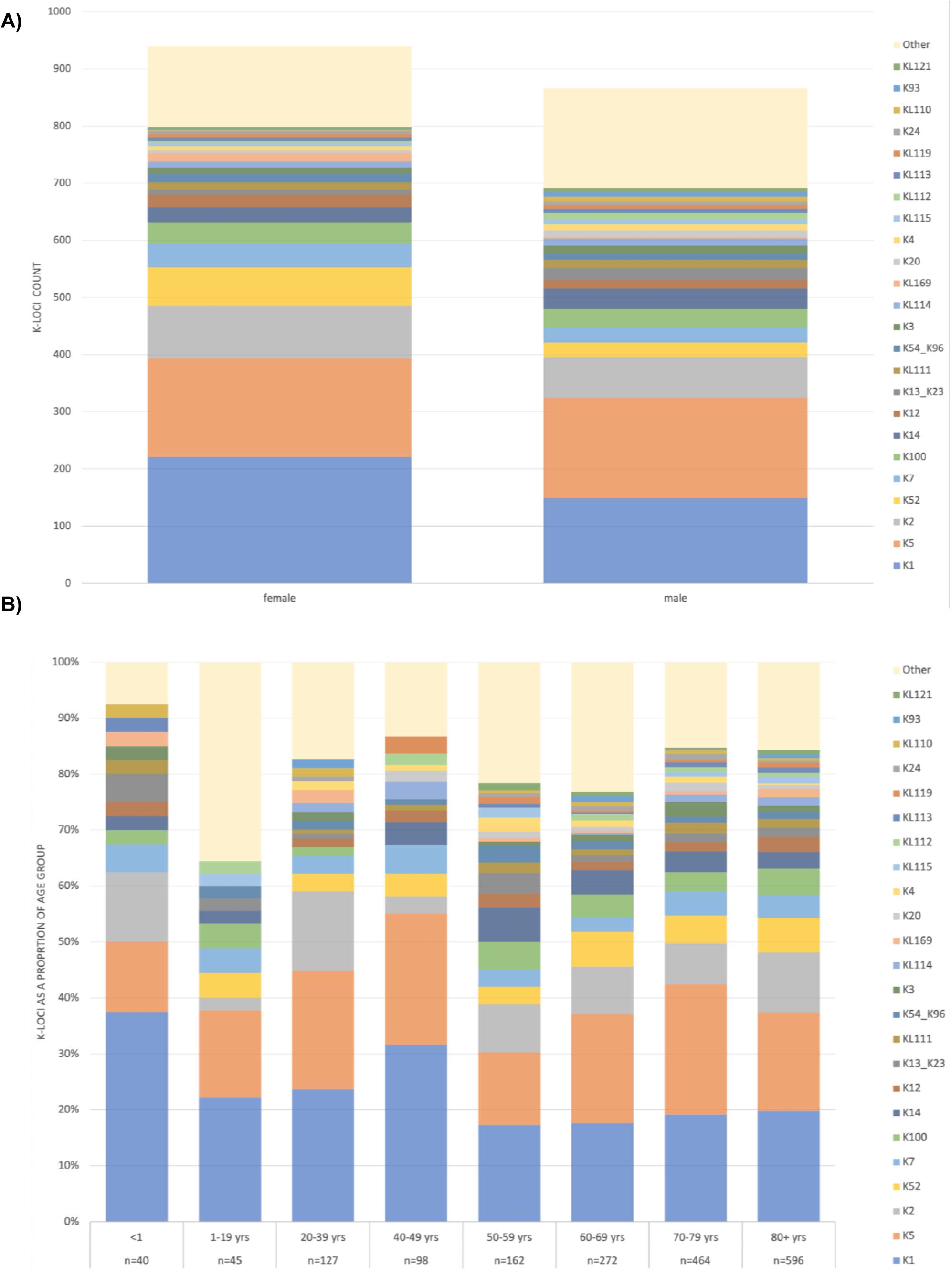
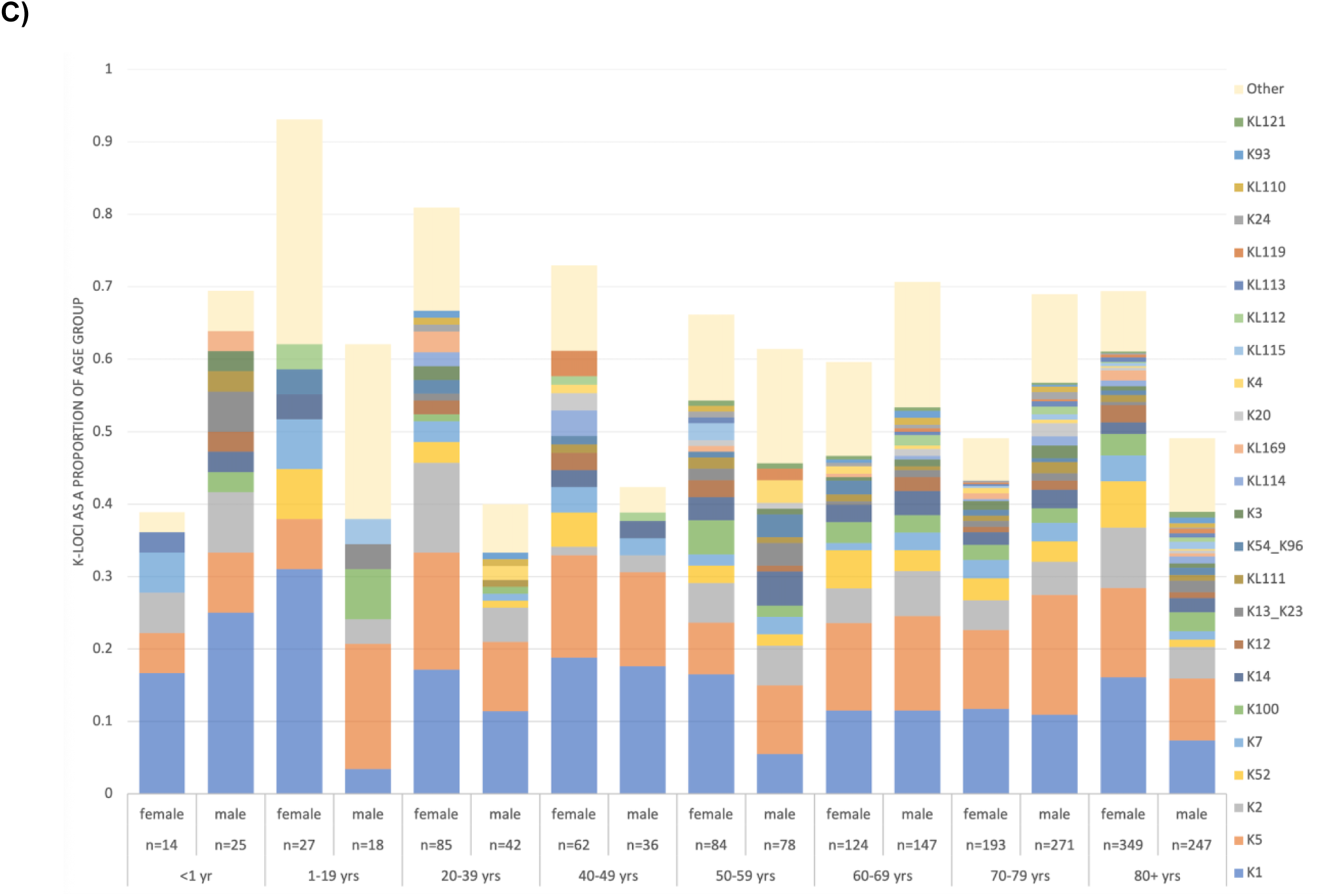
K-loci in UK BSIs A) K-loci by sex B) K-loci per age group C) K-loci per age group by sex.

**Supplementary Figure 2.**
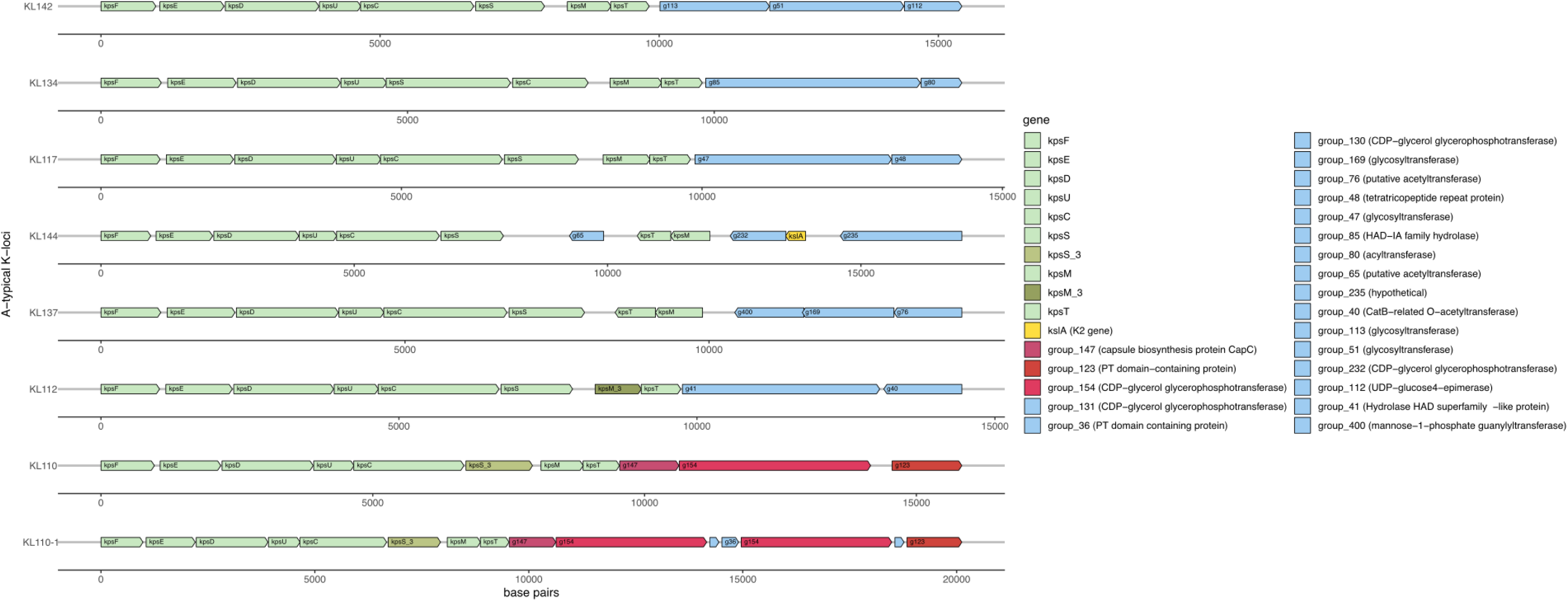
Genetic architecture of atypical G2 K-loci. The conserved *kps* Regions 1 and 3 genes are light green, with rarer gene clusters in darker green. Region 2 genes observed in known K-types are in yellow, and those shared between atypical K-loci are in red. Genes only seen in one locus are blue. The top *E. coli* blast match is shown in brackets for unannotated gene clusters.

**Supplementary Figure 3.**
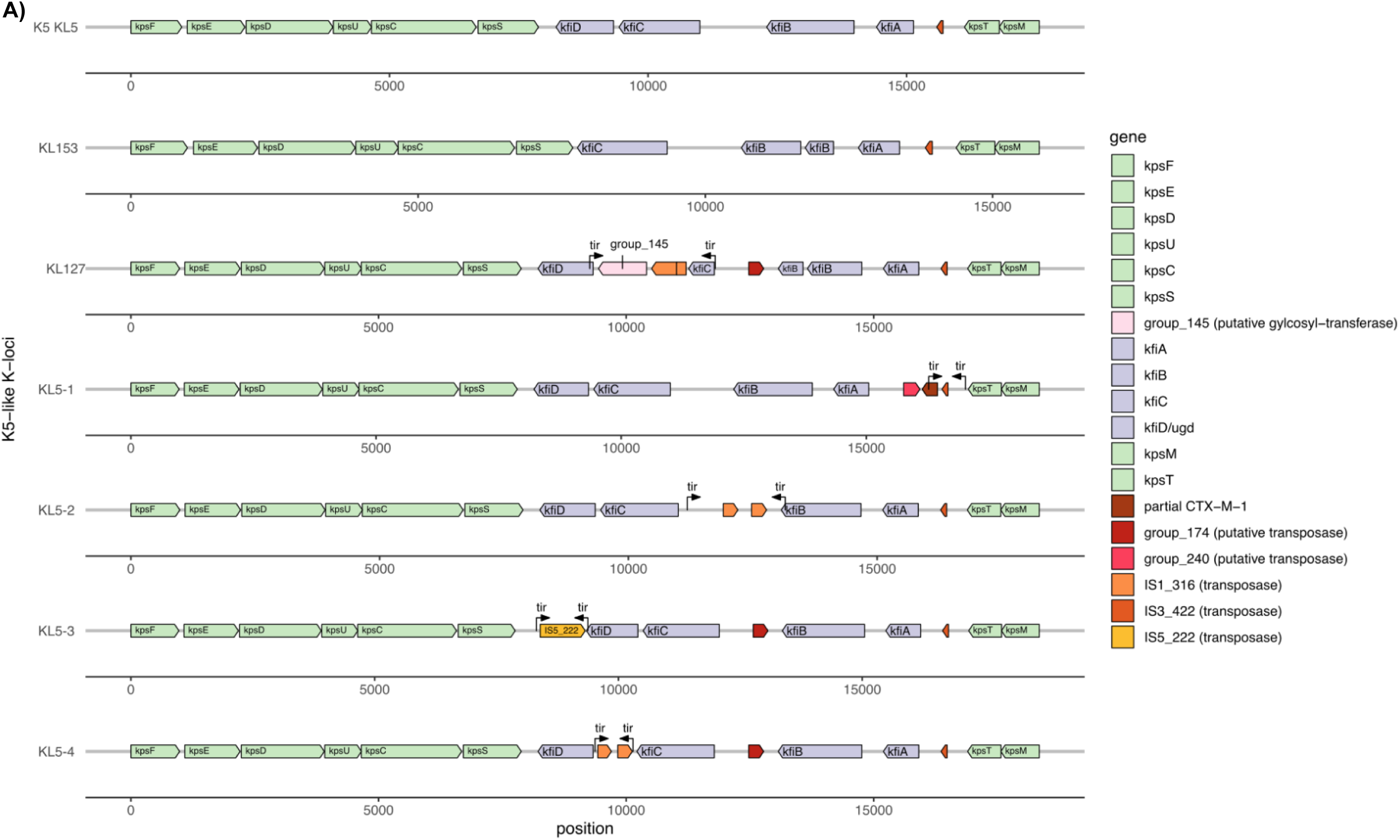
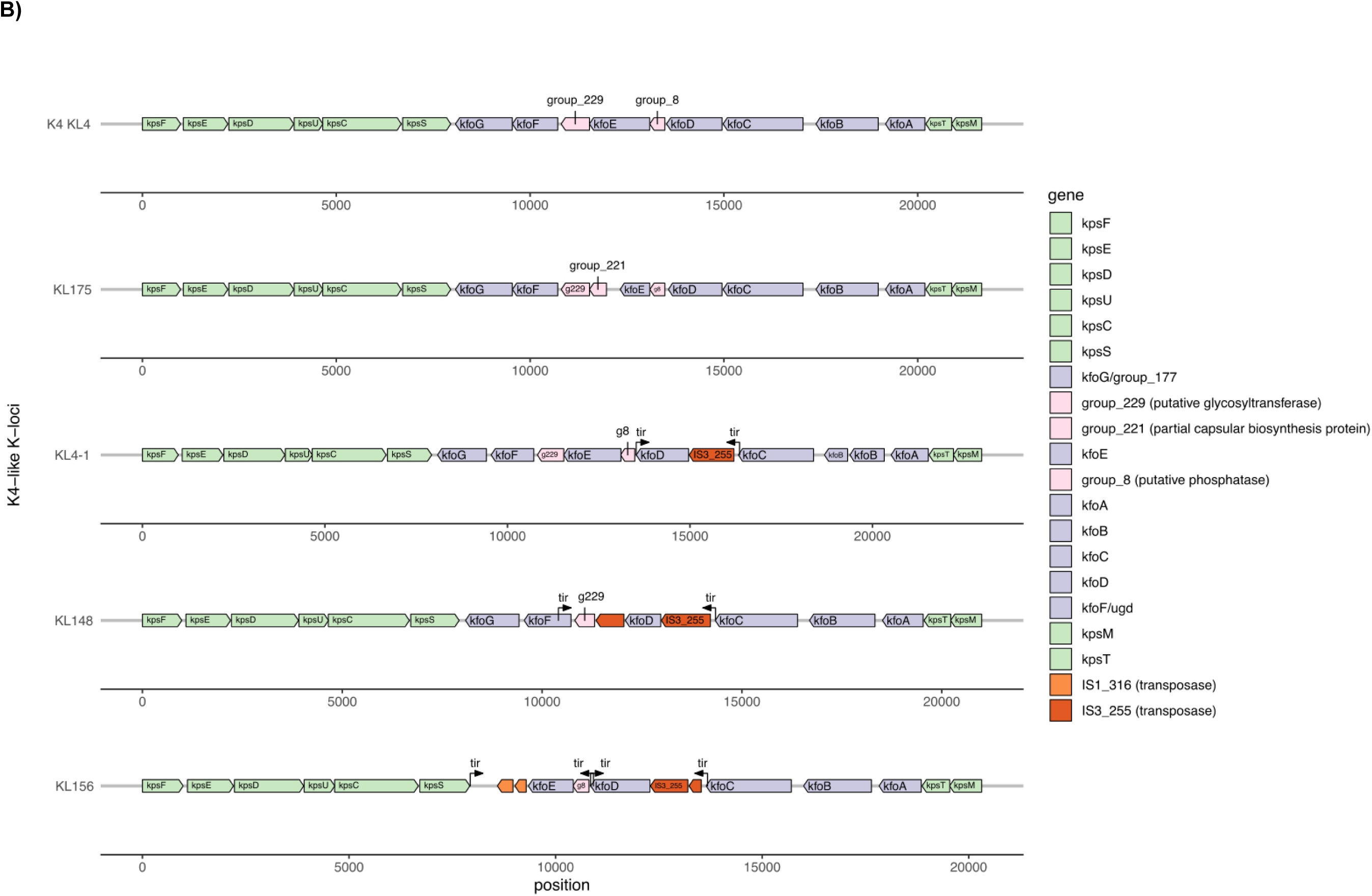
Gene presence and absence for A) the K5-related loci and B) the K4-related loci. Region 1 and 3 *kpsF-kpsM* (green), Region 2 containing genes determining the capsular type (purple), and putative capsular genes (pink). IS genes and other non-capsular genes are included here (other). When the capsular-specific gene name in the literature applies to a gene found across multiple capsular types in the database, the generic gene cluster name is also provided. Putative and known functions are shown in brackets.

